# Validation of a Pancreatic Cancer Detection Test in New-Onset Diabetes Using Cell-Free DNA 5-Hydroxymethylation Signatures

**DOI:** 10.1101/2021.12.27.21268450

**Authors:** David Haan, Anna Bergamaschi, Gulfem D Guler, Verena Friedl, Yuhong Ning, Roman Reggiardo, Michael Kesling, Micah Collins, Bill Gibb, Adriana Pitea, Kyle Hazen, Steve Bates, Michael Antoine, Carolina Fraire, Vanessa Lopez, Roger Malta, Maryam Nabiyouni, Albert Nguyen, Tierney Phillips, Michael Riviere, Aaron Scott, Eric Nilson, Judy Sheard, Melissa Peters, Shimul Chowdhury, Wayne Volkmuth, Samuel Levy

## Abstract

**BACKGROUND:** Pancreatic cancer (PaC) has poor (10%) 5-year overall survival, largely due to predominant late-stage diagnosis. Patients with new-onset diabetes (NOD) are at a six-to eightfold increased risk for PaC. We developed a pancreatic cancer detection test for the use in a clinical setting that employs a logistic regression model based on 5-hydroxymethylcytosine (5hmC) profiling of cell-free DNA (cfDNA).

**METHODS:** cfDNA was isolated from plasma from 89 subjects with PaC and 596 case-control non-cancer subjects, and 5hmC libraries were generated and sequenced. These data coupled with machine-learning, were used to generate a predictive model for PaC detection, which was independently validated on 79 subjects with PaC, 163 non-cancer subjects, and 506 patients with non-PaC cancers.

**RESULTS:** The area under the receiver operating characteristic curve for PaC classification was 0.93 across the training data. Training sensitivity was 58.4% (95% confidence interval [CI]: 47.5– 68.6) after setting a classification probability threshold that resulted in 98% (95% CI: 96.5–99) specificity. The independent validation dataset sensitivity and specificity were 51.9% (95% CI: 40.4–63.3) and 100.0% (95% CI: 97.8–100.0), respectively. Early-stage (stage 1 and 2) PaC detection was 47.6% (95% CI: 23%–58%) and 39.4% (95% CI: 32%–64%) in the training and independent validation datasets, respectively. Sensitivity and specificity in NOD patients were 55.2% [95% CI: 35.7–73.6] and 98.4% [95% CI: 91.3–100.0], respectively. The PaC signal was identified in intraductal papillary mucinous neoplasm (64%), pancreatitis (56%), and non-PaC cancers (17%).

**CONCLUSIONS:** The pancreatic cancer detection assay showed robust performance in the tested cohorts and carries the promise of becoming an essential clinical tool to enable early detection in high-risk NOD patients.

## Introduction

Pancreatic cancer is the third-leading cause of cancer death in the United States.^1^ To date, there are no early detection tools available to patients. It is estimated that 60,430 new pancreatic cancer diagnoses will be made in the United States in 2021, and the annual incidence is predicted to increase to 93,000 by 2040.^2^ Pancreatic cancer has the poorest overall survival of all the major cancer types, with a 5-year relative survival rate of 10%, which has remained relatively unchanged for more than 40 years. Poor survival outcomes for patients with pancreatic cancer are attributable, largely, to the late-stage diagnosis of the disease in the majority (82%) of patients, including distant metastasis in 52% of patients.^1^ Late diagnosis deprives patients of potentially curative treatments, such as surgery,^3^ and negatively impacts survival rates, with 5-year survival rates reduced more than 10-fold from 39% to 3% for localized compared with distant metastatic disease.^1^ Notably, the 5-year survival rate for stage IA (node-negative pancreatic cancer <2 cm) exceeds 80%.^4^ It is therefore evident that early diagnosis is paramount for better survival outcomes in patients with pancreatic cancer.

The lack of symptoms in early-stage pancreatic cancer indicates a need for screening in asymptomatic individuals, but, as a result of the low prevalence of pancreatic cancer,^1^ the United States Preventive Services Task Force concluded that the potential benefits of screening for the disease in asymptomatic adults does not outweigh the potential harm due to overdiagnosis and overtreatment.^5^ There is, however, evidence that higher pancreatic cancer risk groups with increased incidence rates may benefit from screening.^6^

While there are certain genetic syndromes with increased risk for pancreatic cancer (e.g., Peutz-Jeghers syndrome or germline mutations in the *CDKN2A, BRCA2*, or *PALB2*), they account for only a small portion (10%–15%) of cases.^7,8^ The majority of cases emerge sporadically in patients with no familial history of pancreatic cancer. Risk factors for pancreatic cancer include age, with pancreatic cancer incidence peaking at 65–69 years in males and 75–79 years in females,^9^ and a personal history of chronic pancreatitis,^10^ intraductal papillary mucinous neoplasm (IPMN),^11^ gallstone disease,^12^ and pancreatic cysts.^13^ Other well-recognized risk factors for pancreatic cancer include a personal history of cigarette smoking,^14^ alcohol consumption,^15^ and obesity.^16^ Of note, more than 50% of patients diagnosed with pancreatic cancer have a prior diagnosis of diabetes mellitus.^17^ Furthermore, while patients with diabetes mellitus have a 1.5-to 2-fold increased risk of developing pancreatic cancer compared with the general population,^18^ the risk is increased six-to eightfold in patients ≥50 years of age with new-onset (≤36 months previously) diabetes (NOD).^19^ Indeed, nearly 25% of all new diagnoses of pancreatic cancer in the United States are identified in patients with NOD.^20^

The potential risks for overdiagnosis and overtreatment due to pancreatic cancer screening can be mitigated by employing new technologies with high specificity and sensitivity in the appropriate high-risk population enriched for disease incidence, such as patients with NOD.^21^ Surveillance of the NOD population for pancreatic cancer therefore presents an opportunity to shift pancreatic cancer diagnosis to earlier-stage disease and to improve outcomes with timely intervention. Currently, the diagnosis of pancreatic cancer relies on non-invasive imaging methods, such as computerized tomography (CT) and magnetic resonance imaging, as well as invasive imaging methods, including endoscopic ultrasound (EUS).^13^ These methods are not amenable to surveillance of high-risk individuals due to various limitations, including exposure to ionizing radiation of CT scans and complications associated with invasive methodology, such as EUS.^22,23^ However, evidence from the Cancer of the Pancreas Screening trial, which is conducted by the National Cancer Institute, Johns Hopkins University and others, suggests that pancreatic cancers can be detected earlier and have better prognosis when screened.^24^ It has been proposed that serological biomarkers that enrich the NOD population further to identify patients at higher risk of developing pancreatic cancer could be useful for early detection of pancreatic cancer.^21^

An approved, non-invasive, primary screening test for pancreatic cancer detection based on molecular subtyping using mutation analysis or gene expression signatures has, so far, proved elusive.^25^ Epigenetic control of DNA state and chromatin regulation is known to underpin cancer onset and progression,^26,27^ and interest in epigenetic subtyping for tumor detection and characterization has increased in recent years.^28^ Screening tests based on epigenetic profiling have recently been approved for cancers, including colorectal, breast, and lung cancers.^28^

5-hydroxymethylcytosine (5hmC) is a stable epigenetic mark that arises as the first step of active demethylation of the cytosine base in DNA by ten-eleven translocation enzymes, marking regions of active transcription and gene regulation.^29^ 5hmC is positively correlated with gene expression and regulation in multiple biological contexts.^30-32^ As such, 5hmC profiles have yielded distinctive signatures that enable definition of tissue identity and cellular states,^31,33^ and 5hmC is a valuable marker to identify the tissue of origin. Notably, 5hmC profiles can also be used to detect cancer in cell-free DNA (cfDNA) in the plasma of patients with cancers, including pancreatic, lung, hepatocellular, colon, and gastric cancers.^34-36^

This paper describes the development and validation of a 5hmC based, machine learning– enabled assay, suitable for clinical setting for the detection of pancreatic cancer signatures using a single blood draw. The test is intended to be offered to individuals at higher risk for pancreatic cancer such as newly diagnosed diabetics.

## Methods

### Study design and cohorts

The non-invasive liquid biopsy analysis of epigenomic signatures in multiple cancer types, which was a case-control study with longitudinal follow up for non-cancer participants (NCT03869814), enrolled subjects with and without cancer, including pancreatic cancer, from 146 sites across the United States. All participants provided informed consent, and the study was approved by the Institutional Review Boards (IRBs) responsible at each site (Sterling IRB or WIRB–Copernicus Group, Inc.). The study protocol submission, IRB approval, and specimen handling across all sites were managed by several contract research organizations

Male and female participants aged between 45 and 75 years were enrolled in the study. Exclusion criteria included blood product transfusion, surgery or invasive procedure requiring general anesthesia within 1 month of enrollment, organ transplant or major trauma within 6 months of enrollment; previous treatment with molecularly targeted immune modulation therapy within 12 months of enrollment; or current or previous pregnancy within 12 months of enrollment. Study control subjects enrolled in the pancreatic cancer cohort were required to have no prior history of cancer and a pathologically confirmed pancreatic cancer diagnosis. Additionally, participants in the pancreatic cancer cohort were required to be cancer treatment–naive at the time of study enrollment and blood sample acquisition.

The case-control study was divided into two prespecified datasets, and the cohort samples were collected and divided in two groups: a training dataset and an independent validation dataset. Samples for training predictive, logistic regression-based models were obtained from participants within the training cohort who were negative for IPMN, NOD, pancreatitis, and cancer of any type other than pancreatic.

### Sample Collection and Plasma Preparation

Plasma was isolated from whole blood specimens obtained by routine venous phlebotomy at the time of enrollment. Whole blood (2×10 mL) was collected in Cell-Free DNA BCT® tubes (Streck, La Vista, NE) per manufacturer’ s protocol and maintained at 15°–25°C before processing within 24–48 hours of venipuncture. To separate plasma, tubes were centrifuged at 1600×g for 10 minutes, and the plasma was transferred to new tubes for further centrifugation at 16,000×g for 10 minutes. The final plasma was aliquoted for frozen storage at −80°C before cfDNA isolation.

### cfDNA Isolation

cfDNA isolation was carried out using the MagMax™ cfDNA isolation kit (Thermo Fisher Scientific, Waltham, MA) according to the manufacturer’ s protocol using an automated liquid handler (Hamilton Company, Reno, NV). Isolated cfDNA was quantified using Quant-iT™PicoGreen™ (Thermo Fisher Scientific, Waltham, MA) and stored at -20°C until required.

### 5hmC Assay Enrichment

Isolated cfDNA from a single BCT was normalized to 10 ng in a 96-well-plate using an automated liquid handler (Beckman Coulter Life Sciences, Indianapolis, IN) and ligated to sequencing adapters. 5hmC bases were biotinylated via a two-step chemistry and enriched by binding to Dynabeads M-270 Streptavidin (Thermo Fisher Scientific, Waltham, MA). Following the enrichment of 5hmC fragments, libraries were amplified by polymerase chain reaction and normalized to 1 ng/uL using an automated liquid handler (Hamilton Company, Reno, NV). After normalization, libraries were pooled and sequenced on a NovaSeq sequencer (Illumina Inc., San Diego, CA).

### Model training

Sequencing data were collated using NovaSeq Control Software v1.7.0 (Illumina Inc., San Diego, CA). Raw data processing and demultiplexing were performed using bcl2fastq conversion software (Illumina Inc., San Diego, CA) to generate sample-specific FASTQ output. Sequencing reads were analyzed by a computational pipeline implemented as a Nextflow script, which aligns the reads to the human genome build 38 reference genome using the BWA-MEM2 algorithm. Metrics were computed by the pipeline via Picard and additional programs to assess the quality of the sequencing data. Samples passing quality control metrics were scored by the pipeline using a a logistic regression model trained on case/control data. The machine-learning classification algorithm was trained as follows: each sample included in the training dataset was analyzed with the bioinformatics pipeline as described above. The pipeline divided the genome into functional regions pertaining to annotated gene bodies from Gencode human annotation version 31 (GRCh38.p12) and then counted the number of 5hmC library read pairs mapped to each gene body region correcting for differences in coverage using counts per million (CPM) mapped reads. An elastic net logistic regression model was built using the R package glmnet using the CPM output by the pipeline as the feature vector, with elastic net mixing ratio α and the regularization parameter λ optimized using fivefold cross-validation. The final model contained 100 non-zero coefficients, resulting in an easily interpretable classification model. To classify each sample as cancer and non-cancer, a probability threshold was determined that resulted in 98% specificity of the non-cancers in the training data (Figure 1).

**Figure 1.**
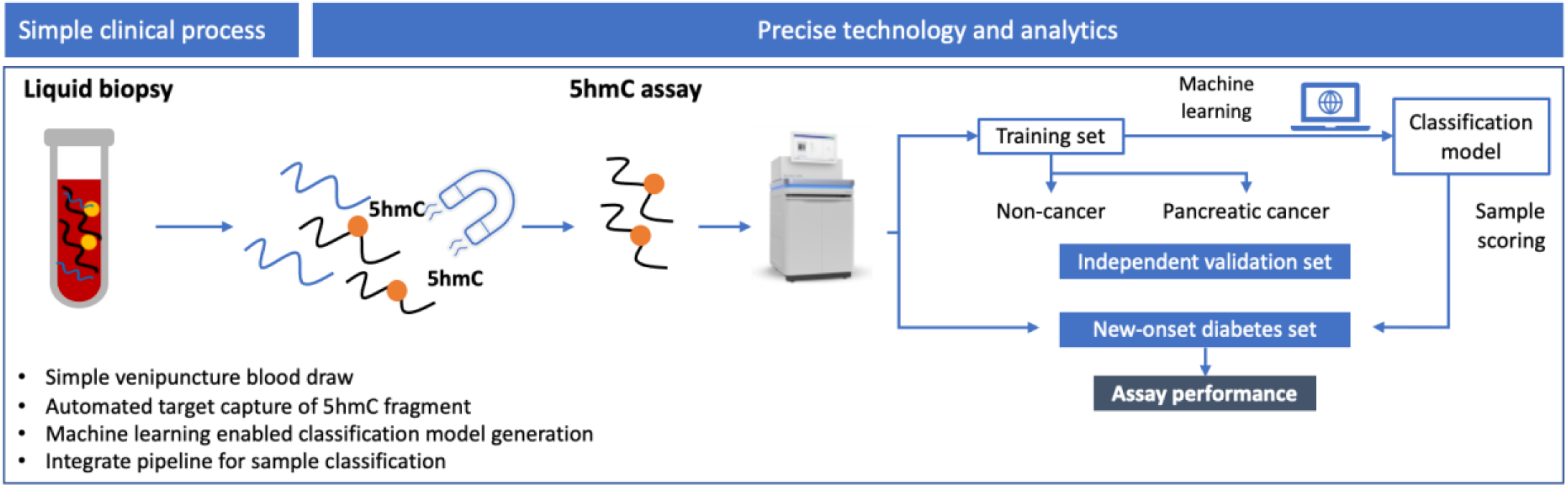
5hmC enrichment and machine learning workflow to develop and validate a predictive model for detection of pancreatic cancer in patients with NOD.

### Model Validation

The fully trained model was integrated into our automated computational pipeline as a Docker container executed with a Nextflow pipeline implemented on Amazon Web Service (Amazon, Seattle, WA). Upon full integration, predictive scores and cancer classification were generated for each sample in the independent validation study. Scoring and cancer classification were performed in a blind fashion and sample labels were revealed after scoring was completed.

### Differential Gene Representation Analysis

For differential gene analysis, we first removed weakly represented genes, we excluded genes that did not have greater than 3 counts per million reads in at least 20 samples. This filter excludes roughly 7.4% of the genes. R package “edgeR”^37^ was employed for differential analysis as follows: (i) generation of a design matrix to compare PaC versus non-cancer cohorts, (ii) removal of heteroscedasticity from the data, (iii) fitting regression models for the comparison of interest, PaC vs non-cancer, (iv) evaluation of the number of differentially represented genes. For the identification of the significantly differentially represented genes, we used the method of Benjamini and Hochberg^38^ to obtain *p*-values adjusted for multiple comparisons. In this report, we use adjusted *p*-value and false discovery rate (FDR) interchangeably. The same process was repeated for NOD samples.

## Results

### Study Cohorts

The training dataset was based on results from 740 plasma samples from 92 participants with pancreatic cancer and 648 without cancer. Of these, three samples from participants with pancreatic cancer and 52 non-cancer samples failed quality control (QC). Most of the QC failures were due to insufficient amount of cfDNA. The final training dataset was based on samples from 89 pancreatic cancer and 596 non-cancer participants (**Table 1** and **Figure 2A**

**Table 1.** Participant demographic characteristics and disease status.

| Variables | Training set |  | Independent Validation set |  |  |
| --- | --- | --- | --- | --- | --- |
|  | Non-cancer<br>(N = 596) | PaC<br>(N = 89) | Non-cancer<br>(N = 163) | PaC<br>(N = 79) | OOPC<br>(N = 506) |
| Age (mean years) | 62 | 67 | 61.0 | 65.3 | 63.6 |
| Male (%) | 36.2 | 56.2 | 62.6 | 48.1 | 47 |
| Stage (%) |  |  |  |  |  |
| I | - | 18.0 | - | 19.0 | 22.3 |
| II | - | 29.2 | - | 22.8 | 20.0 |
| III | - | 10.1 | - | 11.4 | 26.7 |
| IV | - | 40.5 | - | 45.6 | 29.9 |
| NA | - | 2.2 | - | 1.2 | 1.1 |

**Figure 2.**
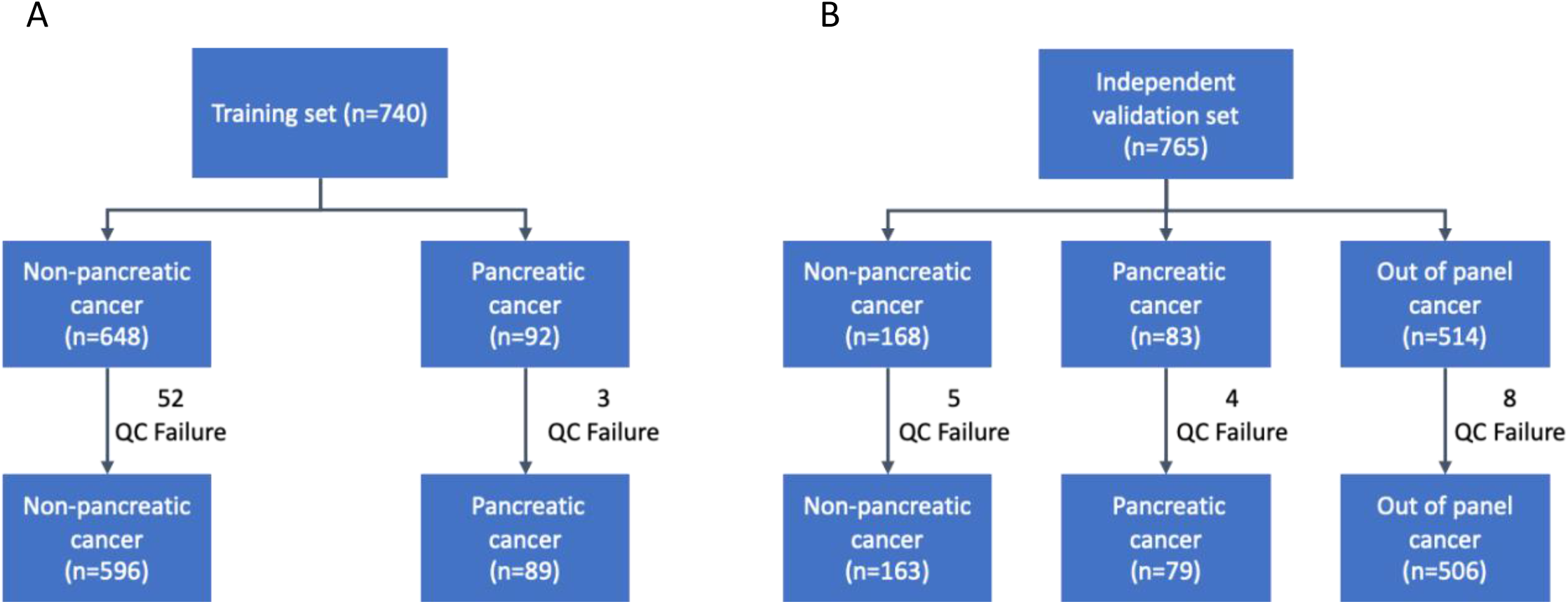
Clinical cohorts for (A) model training and (B) validation.

Samples from 83 participants with pancreatic cancer, 168 participants without cancer, and 514 participants with a cancer diagnosis other than pancreatic cancer (out-of-panel cancer [OOPC]) were utilized for independent validation of the model (**Table 1**). Of these, 79, 163, and 506 samples from participants with pancreatic cancer, without cancer and OOPCs, respectively, successfully completed the QC stage (**Figure 2B; Table 1**). Samples from participants with OOPC included the following cancer types: breast (n = 97), colorectal (n = 146), prostate (n = 35), and lung (n = 228) cancers, were included in the validation.

### Logistic Regression Model Construction and Assay Performance Evaluation

In order to build a discriminator to detect pancreatic cancer from 5hmC profiles taken from cfDNA of patients with and without disease, we employed two distinct cohorts; 89 pancreatic cancer patients and 596 non-cancer subjects from the training dataset. The prediction model was constructed using elastic net logistic regression and built using the fraction of 5hmC counts on each gene as generated by the computational pipeline as a feature vector. The model constructed was optimized via fivefold cross-validation of the elastic net mixing ratio α and the regularization parameter λ. This approach yielded a model that performed with an overall area under the receiver operating characteristic curve (auROC) of 0.93 (**Figure 3A**). Consistent high performance was observed with early-stage disease, with auROCs of 0.87 and 0.91 for stage 1 and 2 diseases respectively, and a range 0.87-0.96 across all disease stages (I-IV) (**Figure 3B**).

**Figure 3.**
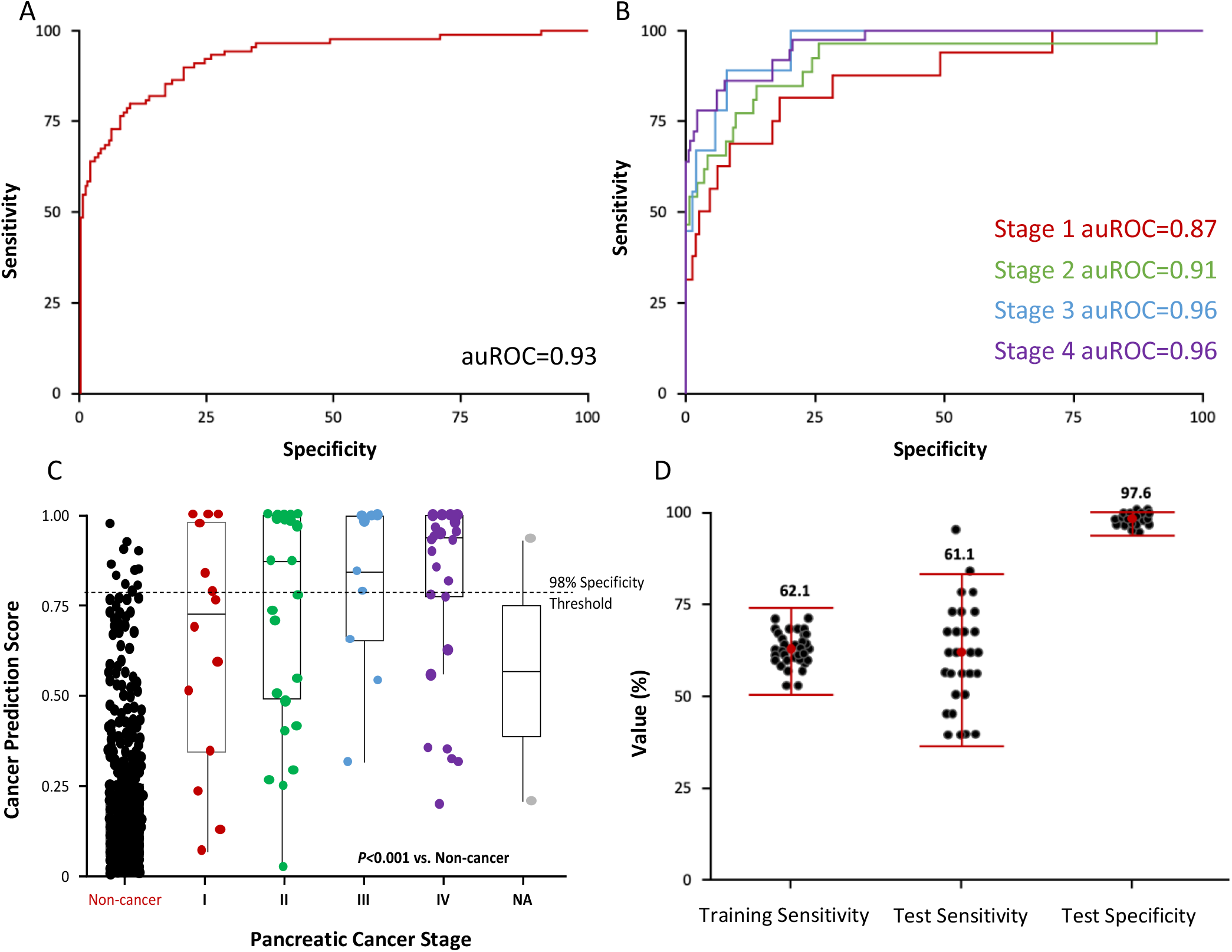
Model training. (A) Predictive modeling using regularized regression model (elastic net) on the training dataset with 89 pancreatic cancer and 596 non-cancer cfDNA. (B) Predictive modeling using regularized regression model (elastic net) on the training dataset. (C) Pancreatic cancer prediction score by disease stage. Dashed line indicates 98% specificity threshold. (D) Fifty iterations of training/test splits of the training dataset showing consistent sensitivity and specificity. 96% of the iterations (black dots) for test sensitivity and specificity fell between the 95% binomial confidence intervals.

The cancer prediction scores, which approached 0 and 1 for non-cancer and pancreatic cancer respectively, showed significant differentiation in scoring between non-cancer and early-stage pancreatic cancer (*P* <0.0001 for both stage 1 and stage 2 vs non-cancer **Figure 3C**). Next, we examined the consistency of the model performance, by generating 50 independent test simulations. Each simulation produced a distinct logistic regression model by randomizing 80:20 (training: test) sample partitions, and the resulting prediction performance across 50 independent tests was highly consistent. Indeed, all 50 simulations produced sensitivity and specificity values for each test within the 95% confidence intervals (CIs), and for both the test sensitivities and specificities, 96% of the values fell within these CIs (**Figure 3D**). After employing a probability threshold to generate a 98% training specificity, the corresponding training and test mean sensitivities and test mean specificity were 62.1% (95% CI, 49.8%– 73.4%), 61.1% (95% CI, 35.7%–82.7%), and 97.5% (95% CI, 92.9%–99.5%), respectively.

### Performance of the Logistic Regression Model in an Independent Validation Cohort

In order to assess how well the constructed pancreatic cancer discriminator was able to detect cancer in a new dataset, a separate study was performed on a set of new patient samples. This independent validation cohort consistent of PaC, non-cancer, and OOPC patient samples processed independently of the training set (**Table 1**), was used to validate the model performance. The samples were scored using the logistic regression model built from training data and using a process that blinded disease labels. The sensitivities and specificities of the training and independent validation datasets are shown in **Table 2A**.

**Table 2.**
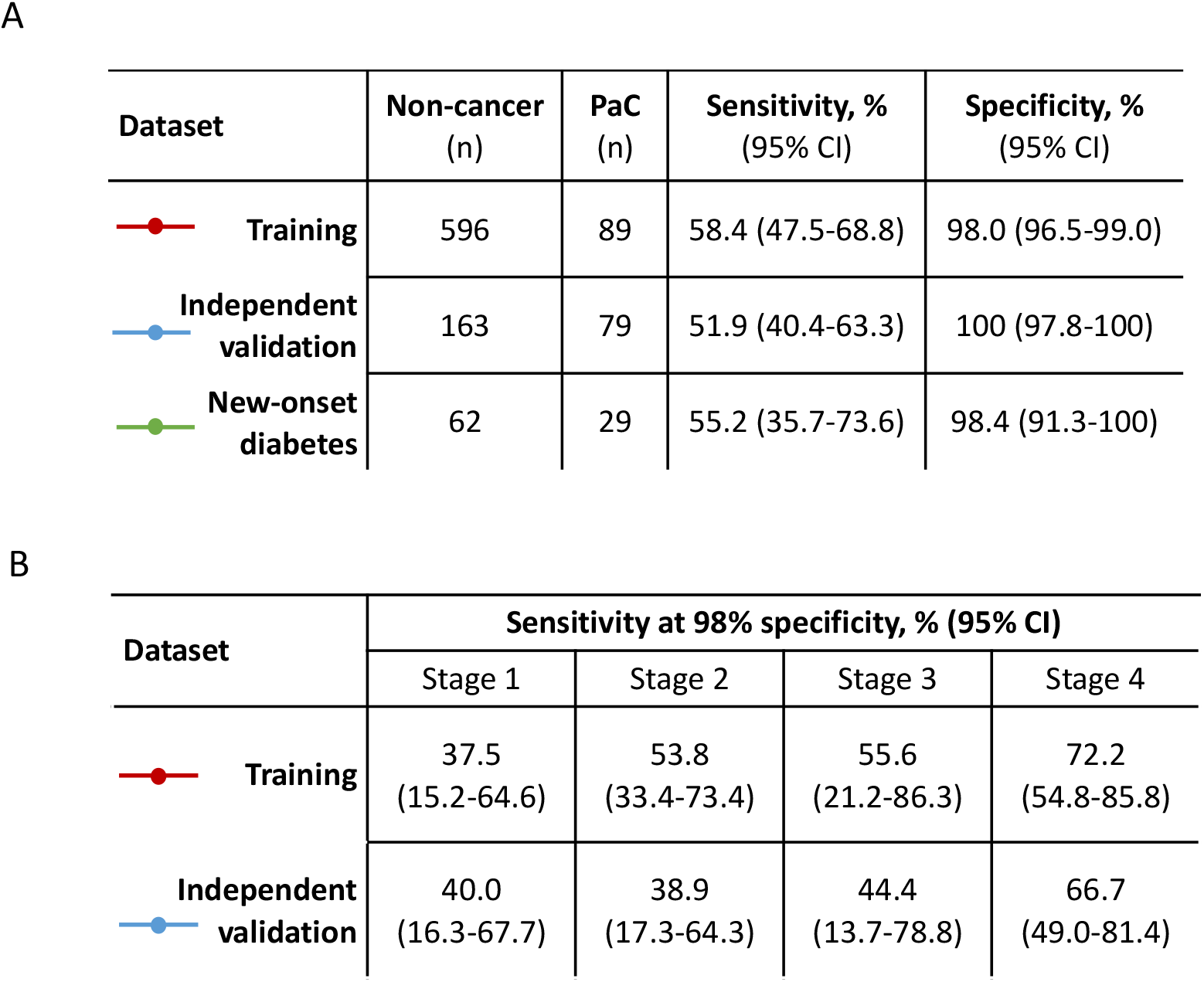
(A) Summary of model performance in the training, independent validation, and NOD datasets. (B) Sensitivity observed by pancreatic cancer stage in the training and independent validation datasets.

| Dataset | Non-cancer<br>(n) | PaC<br>(n) | Sensitivity, %<br>(95% CI) | Specificity, %<br>(95% CI) |
| --- | --- | --- | --- | --- |
| Training | 596 | 89 | 58.4 (47.5-68.8) | 98.0 (96.5-99.0) |
| Independent<br>validation | 163 | 79 | 51.9 (40.4-63.3) | 100 (97.8-100) |
| New-onset<br>diabetes | 62 | 29 | 55.2 (35.7-73.6) | 98.4 (91.3-100) |

B
| Dataset | Sensitivity at 98% specificity, % (95% CI) |  |  |  |
| --- | --- | --- | --- | --- |
|  | Stage 1 | Stage 2 | Stage 3 | Stage 4 |
| Training | 37.5<br>(15.2-64.6) | 53.8<br>(33.4-73.4) | 55.6<br>(21.2-86.3) | 72.2<br>(54.8-85.8) |
| Independent<br>validation | 40.0<br>(16.3-67.7) | 38.9<br>(17.3-64.3) | 44.4<br>(13.7-78.8) | 66.7<br>(49.0-81.4) |

These datasets were also used to examine the model’ s performance in detecting pancreatic cancers at different disease stages. A sensitivity of 47.6% (CI: 23%-58%) and 39.4% (CI: 32%-64%) was observed in early-stage disease (stage I and II samples combined) in the training and independent validation datasets, respectively (**Table 2B**). Next we examined the distribution of the cancer prediction scores, which validated the findings from the training set and confirmed a significant difference in score between non-cancer and early-stage pancreatic cancer (*P* <0.0001 for both stage I and stage II vs non-cancer; **Figure 4**).

**Figure 4.**
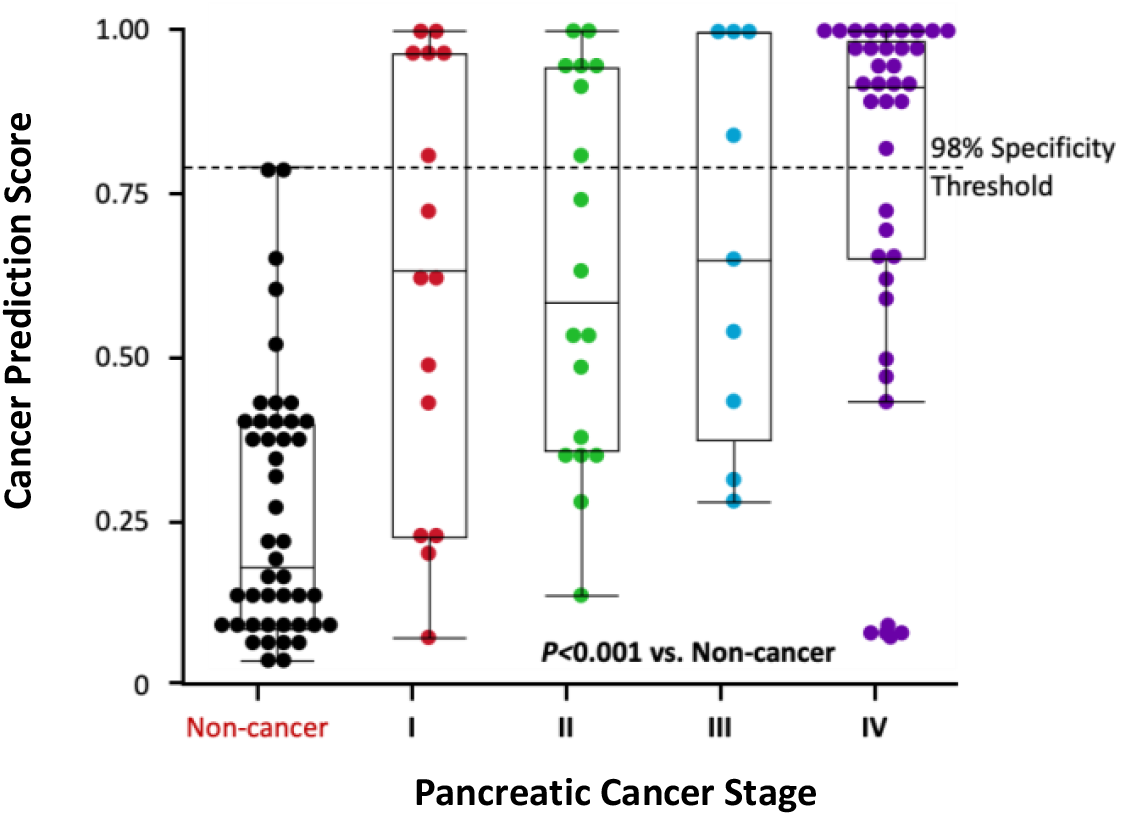
Model validation. Pancreatic cancer prediction score disctribution in the independent validation set stratified by disease stage. Dashed line indicates 98% specificity threshold.

Next, model performance was examined in a set of NOD subjects. The prediction performance in the NOD cohort was similar to that of the non-NOD cohort, with sensitivity and specificity of 55.2% (95% CI, 35.7–73.6) and 98.4% (95% CI, 91.3–100.0), respectively (**Table 2A**), suggesting that the model detects a pancreatic cancer signal independently of diabetes status.

When applied to samples from participants with no evidence of any cancer but with IPMN or pancreatitis, the model predicted pancreatic cancer in 21 of 33 (64%) participants with IPMN and five of nine (56%) participants with pancreatitis (**Figure 5**). Of note, the majority of IPMNs that were classified as pancreatic cancers, had moderate to high dysplasia and 1 case presented additional pancreatic intraepithelial neoplasia foci (PanIN1) and a second case with PanIN2.

**Figure 5.**
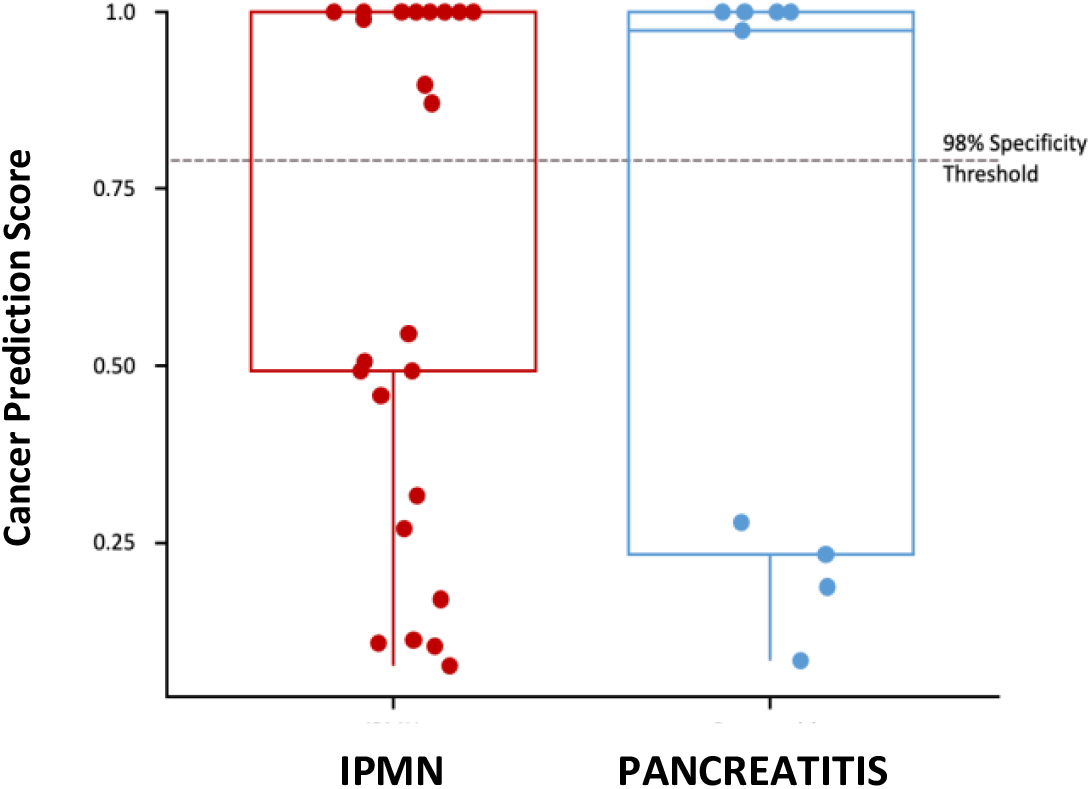
Test performance in pancreatitis and IPMN samples. Cancer prediction scores for cfDNA from patients with IPMN or pancreatitis but no evidence of cancer. Dashed line represents the probability threshold for classification as pancreatic cancer; 21 of 33 and 5 of 9 patients with IPMN and pancreatitis were classified as having pancreatic cancer.

In order to assess the false detection rate for non-PaC cancer the logistic regression model was applied to OOPCs in the independent validation set, containing breast, colorectal, lung, and prostate cancers (**Table 3**). Not surprising, gastrointestinal cancer such as colorectal, had a higher rate of misclassification compared with lung, prostate and breast cancers (23.2% vs 17.1% 11.4% and 10.3%, respectively). After adjusting for cancer incidence rates, the impact of these OOPC false positive rates on specificity was only a decrease of 0.34%.

**Table 3.** Out-of-panel cancers test

| ***Cancer type*** | ***Specimens tested (N)*** | ***Positive tests (N)*** | ***Positive tests (%)*** | ***Cancer incidence per 10,000 in >50 yo (N)*** | ***Positives per 10,000 in >50 yo (%)*** |
| --- | --- | --- | --- | --- | --- |
| Breast | 97 | 10 | 10.3 | 73.3 | 7.6 |
| Colorectal | 146 | 34 | 23.2 | 20.0 | 4.7 |
| Lung | 228 | 39 | 17.1 | 33.6 | 5.7 |
| Prostate | 35 | 4 | 11.4 | 11.1 | 1.3 |

The 5hmC preferences in genes from a prior study^36^ were shown be prevalent and pervasive, which lead to the question whether such preferences also occurred in the cfDNA of NOD patients. To this end, differential analysis of 5hmC densities were performed and demonstrated 7,440 hyper-hydroxymethylated and 7,177 hypo-hydroxymethylated (adjusted P value threshold of 0.05 [Benjamini-Hochberg method]) genes in cfDNA from pancreatic cancer patients compared to non-cancer controls (**Figure 6A**), similar to previous findings^36^. The differential hydroxymethylated genes identified in pancreatic cancer patients with NOD were closely correlated with the ones identified in the pancreatic cancer group without NOD (**Figure 6B**). A close correlation was observed between the normalized enrichment scores (NESs) obtained by gene set enrichment analysis for the Molecular Signatures Database (MSigDB) C6 oncogenic pathways comparing pancreatic cancer in unselected patients or pancreatic cancers in patients with NOD to non-cancer controls (**Figure 6C**). Overall, these results demonstrate that the pancreatic cancer signal as detected by 5-hydroxymethylation signatures are similar between NOD and non-NOD populations.

**Figure 6.**
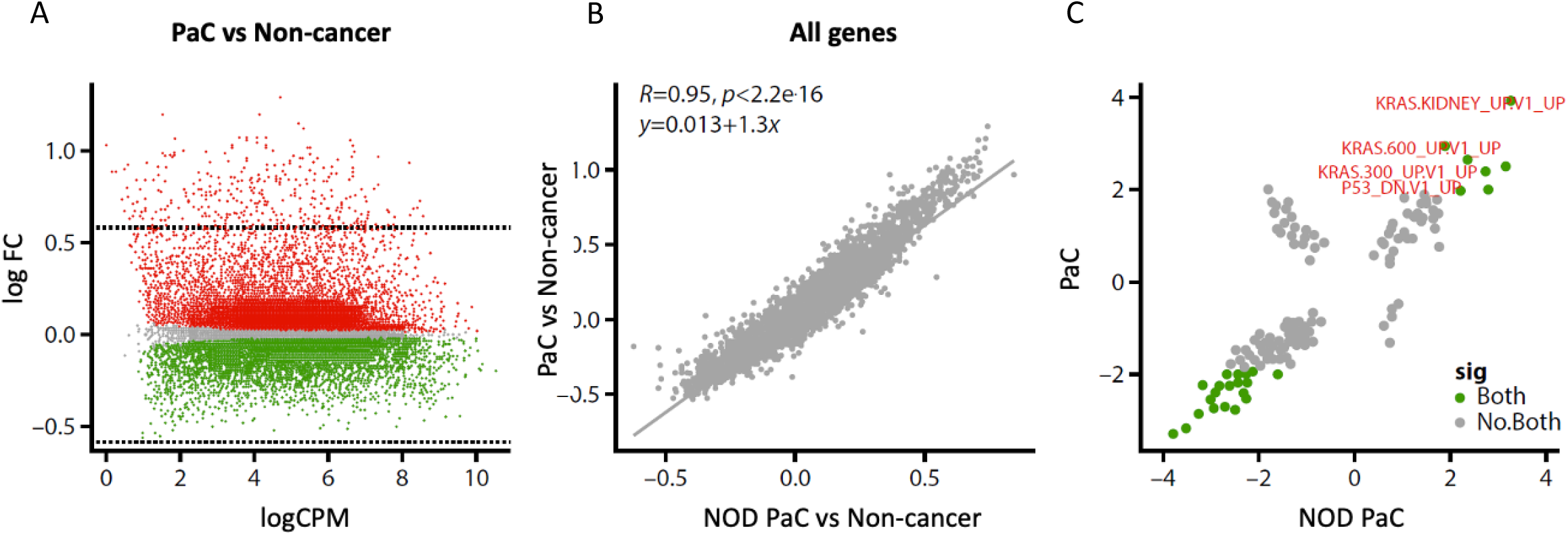
Differential Gene Analysis. 5hmC counts in genes from cfDNA of pancreatic cancer in NOD and non-NOD patients show similar biology. (A) MA-Plot showing all genes with differential 5hmC representation. Red and green, respectively, denote increased or decreased 5hmC density in PaC compared to non-cancer with adjusted *P* value <0.05 (Benjamini– Hochberg). (B) Correlation between fold change of 5hmC representation over all genes comparing PaCs (y-axis) or PaCs with NOD only (x-axis) to non-cancer controls. Pearson correlation as well as *P* values as determined by two-sided *t*-test are reported in the panel. (C) Scatter plot of the NESs obtained by gene set enrichment analysis using MSigDB C6 oncogenic pathways collection comparing PaCs (y-axis) or PaCs with NOD only (x-axis) to non-cancer. Green dots represent significant gene sets enriched in pancreatic cancer with and without NOD. Grey dots represents gene sets non significantly enriched in either cohort.

## Discussion

Pancreatic cancer is a lethal disease that has poorer outcomes when identified at later compared with earlier stages of the disease course.^1^ However, as pancreatic cancer is largely asymptomatic in the earlier stages, it is generally only identified when disease is already advanced.^1,13^ Early identification of pancreatic cancer through screening of the general population is not recommended due to the low incidence of the disease in the general population.^5^ However, targeted screening is recommended for individuals whose lifetime risk of developing pancreatic ductal adenocarcinoma is higher than 5%, which includes individuals with at least two first-degree relatives with the disease, patients with hereditary pancreatitis or particular genetic syndromes, and patients with mucinous cystic lesions of the pancreas.^7^ Other high-risk groups include patients with NOD.^17,19^ A bidirectional and complex association between pancreatic cancer and diabetes mellitus has been documented in many epidemiological studies,^39^ but the exact temporal or causal relationship between diabetes and pancreatic cancer remains undetermined. Accordingly, the relationship between diabetes mellitus and pancreatic cancer has been identified as a research priority in pancreatic cancer research.^40^

Diabetes mellitus is an increasing major public health problem worldwide, and it is estimated that in 2018, 13.0% (34.1 million) US adults had diabetes, increasing to 26.8% in adults ≥65 years.^41^ The prevalence of diabetes is increasing worldwide. It is estimated that in 2060, more than 60 million patients will have been diagnosed with diabetes in the United States alone.^42^ A previous study has demonstrated that the 3-year cumulative incidence rate of pancreatic cancer in patients with NOD is 0.85%,^19^ meaning that nearly 1 in every 100 patients with NOD have a diagnosis of pancreatic cancer within 3 years of diabetes diagnosis. This frequency of pancreatic cancer in NOD patients is similar to that of current colorectal and lung cancer screening programs.^43,44^ In the past decades, there has been much attention on the role of biomarkers in cancer screening, diagnosis, and prognostication.^45^ Elevated concentrations of carbohydrate antigen 19-9 (CA19-9), the only biomarker routinely used in the management of pancreatic cancer, have been shown in samples taken before diagnosis. However, CA19-9 is not recommended for screening because it is not expressed in individuals with a Lewis-negative genotype and is elevated in many other benign and malignant diseases.^46^ Furthermore, pancreatic tumors are highly heterogeneous, both within and between individuals^47^; therefore, single biomarkers are unlikely to have adequate sensitivity for detection. Recently, several studies, including the NOD cohort of the Consortium for the Study of Chronic Pancreatitis, Diabetes, and Pancreatic Cancer, which is designed to recruit 10,000 patients with new-onset of diabetes after age 50 years,^48^ have been launched to obtain samples and monitor patients with a high risk of pancreatic ductal adenocarcinoma (PDAC) to identify early biomarkers for pancreatic cancer.

5hmC-based methods are being increasingly evaluated for the detection of cancer in plasma samples and have been used successfully in lung, hepatocellular, pancreatic,^34,49^ colon, and gastric cancers.^35^ We recently compared 5hmC profiles of plasma-derived cfDNA from case-controlled participants with and without pancreatic cancer and demonstrated differential 5hmC levels for thousands of genes. The most significant increases in 5hmC observed with genes were related to pancreas development (*GATA4, GATA6, PROX1, ONECUT1*, and *MEIS2*) and/or implicated in cancer (*YAP1, TEAD1, PROX1, ONECUT2, ONECUT1*, and *IGF1*).^36^ Gene set enrichment analysis of the genes with the greatest 5hmC changes revealed gene sets that are closely relevant to PaC biology (such as activation of *Kirsten rat sarcoma virus* signaling and downregulation of *p53* signaling), which are known drivers of pancreatic cancer^50,51^ and are associated with survival and recurrence in pancreatic cancer patients.^52,53^ Furthermore, these genes were enriched in targets of transcription factors known to be involved in PaC oncogenesis or metastasis, such as nuclear factor of activated T-cells and forkhead box protein A (hepatocyte nuclear factor 3).^54,55^

Artificial intelligence methodologies have emerged as valuable tools for risk stratification and identification in healthcare and has potential for use in early detection of pancreatic cancer.^56,57^ We have used machine learning to develop a predictive model based on pancreatic cancer 5hmC cfDNA signatures and samples from large cohorts of patients with pancreatic cancer and non-cancer controls. These groups were generally well matched clinical, with a higher proportion of females observed in the control group, likely due to the patient accrual from a simultaneously executed breast cancer study. The test demonstrates good performance in separating pancreatic cancer cfDNA from controls (area under the curve = 0.93) and represents a high-accuracy test for identification of pancreatic cancer in NOD patients. The model yielded a mean test sensitivity of 61.1% (95% CI: 35.7%–82.7%) and a mean test specificity of 97.6% (95% CI: 93%–99.5%) measured across 50 test simulations within the training dataset.

When tested on an independent validation set, including samples from 79 patients with pancreatic cancer (41.8% early stage) and 163 non-cancer controls, the model yielded a classification performance of 51.9% (95% CI: 40.4%–63.3%) sensitivity and 100% (95% CI: 97.8%–100.0%) specificity. Populations with an increased risk of pancreatic cancer, including patients with NOD, are most appropriate for screening.^7,58^ In an assessment exclusively of patients with NOD (N = 91), our model showed a sensitivity of 55.2% (95% CI: 35.7%–73.6%) and 98.4% (95% CI: 91.3%–100.0%) specificity, allowing identification of 16 of the 29 patients with pancreatic cancer and NOD.

While our model was associated with good specificity in the general and NOD populations (98% and 98.4% specificity, respectively), a measurable signal was, unsurprisingly, observed in patients with other solid cancers. This is almost certainly attributable to common pathways of molecular disruption in a wide range of solid cancers.^59^ Notably, the signal was more common with colorectal compared with breast and lung cancers, which is likely attributable to the interconnected molecular subtypes across various gastrointestinal cancers.^60^ While this pattern was not observed in the independent validation cohort, this is likely to be a consequence of the relatively small patient numbers and wide confidence intervals. It is important to note that identification of these cancers would be of benefit in clinical practice and will enable earlier cancer treatment commencement in these patients.

Similarly, the pancreatic cancer model would likely classify patients with pancreatitis and IPMN as those with pancreatic cancer, with 5 of 9 and 21 of 33 cases with pancreatitis and IPMN, respectively. Chronic pancreatitis, a spectrum of persisting fibro-inflammatory disorders of the exocrine pancreas that alters the organ’ s typical structure and functions,^61^ represents one of the highest risk factors for the development of pancreatic cancer.^62^ Inflammatory stimuli may lead to aberrant DNA methylation homeostasis,^63^ and the progressive increase of DNA methylation levels has been described in studies of chronic inflammatory diseases developing into cancer.^64^ In accordance with this hypothesis, the discrimination between pancreatic cancer and non-cancer was less efficient in patients with pancreatitis than in non-cancer controls with or without NOD. However, the methylation profiles of pancreatitis and pancreatic cancer differ,^65^ so it would be feasible to achieve discrimination with model training based on cfDNA from patients with pancreatitis. It should be noted, however, that identification of patients with pancreatitis is a valuable end point that enables patients to receive timely necessary surgical, medical, or supportive interventions.

IPMNs, which are characterized by intraductal papillary proliferation of mucin-producing epithelial cells that exhibit various degrees of dysplasia, are also well known as precursor lesions for pancreatic cancer.^66^ There is evidence that epigenetic changes are present in IPMN and increase progressively with progression to pancreatic cancer.^67^ It is therefore not surprising that the pancreatic cancer model can detect IPMN. Indeed, the IPMN cases that were classified as pancreatic cancers presented moderate to high levels of dysplasia, and although not yet tested, our model could identify IPMN that could develop into pancreatic cancers and need to be closely monitored to determine whether surgical resection is appropriate.^68^

Finally, the close correlation of 5hmC signatures of pancreatic cancer with and without NOD indicate that the biology of pancreatic cancer is similar in the NOD and non-NOD populations and supports the use of our test in patients with NOD.

In summary, we present the development and validation of a technology platform employing 5hmC signal detection coupled with machine learning that can detect pancreatic cancer in patients with NOD. This precise, scalable, cost-effective technology is a promising clinical tool, suited for the early cancer detection of occult pancreatic cancer in those individuals at higher risk such as NOD patients and has the potential to critically improve patient survival outcomes.

## Data Availability

The Fastq files can be made available upon written request for submission to the study institutional review boards at the participating sites.

## Abbreviations used in this paper

5hmC: 5-hydroxymethylcytosine
auROC: area under the receiver operating characteristic curve
CA19-9: carbohydrate antigen 19-9
cfDNA: cell-free DNA
CI: confidence interval
CPM: counts per million
CT: computerized tomography
EUS: endoscopic ultrasound
IPMN: intraductal papillary mucinous neoplasm
IRB: Institutional Review Board
*KRAS*: *Kirsten rat sarcoma virus*
MSigDB: Molecular Signatures Database
NA: not available
NES: normalized enrichment score
NOD: new-onset diabetes
OOPC: out-of- panel cancer
PaC: pancreatic cancer
PDAC: pancreatic ductal adenocarcinoma
QC: quality control
yo: year old

## REFERENCES

1. Siegel RL, Miller KD, Jemal A. Cancer statistics, 2020. CA Cancer J Clin 2020;70:7–30.

2. Rahib L, Wehner MR, Matrisian LM, et al. Estimated projection of US cancer incidence and death to 2040. JAMA Netw Open 2021;4:e214708.

3. Bachmann J, Michalski CW, Martignoni ME, et al. Pancreatic resection for pancreatic cancer. HPB (Oxford) 2006;8:346–351.

4. Blackford AL, Canto MI, Klein AP, et al. Recent trends in the incidence and survival of stage 1A pancreatic cancer: a surveillance, epidemiology, and end results analysis. J Natl Cancer Inst 2020;112:1162–1169.

5. Owens DK, Davidson KW, Krist AH, et al. Screening for pancreatic cancer: US Preventive Services Task Force reaffirmation recommendation statement. JAMA 2019;322:438–444.

6. Lucas AL, Kastrinos F. Screening for pancreatic cancer. JAMA 2019;322:407–408.

7. Canto MI, Harinck F, Hruban RH, et al. International Cancer of the Pancreas Screening (CAPS) Consortium summit on the management of patients with increased risk for familial pancreatic cancer. Gut 2013;62:339–347.

8. Klein AP. Genetic susceptibility to pancreatic cancer. Mol Carcinog 2012;51:14–24.

9. GBD 2017 Pancreatic Cancer Collaborators. The global, regional, and national burden of pancreatic cancer and its attributable risk factors in 195 countries and territories, 1990–2017: a systematic analysis for the Global Burden of Disease Study 2017. Lancet Gastroenterol Hepatol 2019;4:934–947.

10. Raimondi S, Lowenfels AB, Morselli-Labate AM, et al. Pancreatic cancer in chronic pancreatitis; aetiology, incidence, and early detection. Best Pract Res Clin Gastroenterol 2010;24:349–358.

11. Pergolini I, Sahora K, Ferrone CR, et al. Long-term risk of pancreatic malignancy in patients with branch duct intraductal papillary mucinous neoplasm in a referral center. Gastroenterology 2017;153:1284–1294.

12. Nogueira L, Freedman ND, Engels EA, et al. Gallstones, cholecystectomy, and risk of digestive system cancers. Am J Epidemiol 2014;179:731–739.

13. Kamisawa T, Wood LD, Itoi T, et al. Pancreatic cancer. Lancet 2016;388:73–85.

14. Iodice S, Gandini S, Maisonneuve P, et al. Tobacco and the risk of pancreatic cancer: a review and meta-analysis. Langenbecks Arch Surg 2008;393:535–545.

15. Wang YT, Gou YW, Jin WW, et al. Association between alcohol intake and the risk of pancreatic cancer: a dose-response meta-analysis of cohort studies. BMC Cancer 2016;16:212.

16. Arslan AA, Helzlsouer KJ, Kooperberg C, et al. Anthropometric measures, body mass index, and pancreatic cancer: a pooled analysis from the Pancreatic Cancer Cohort Consortium (PanScan). Arch Intern Med 2010;170:791–802.

17. Sah RP, Nagpal SJS, Mukhopadhyay D, et al. New insights into pancreatic cancer-induced paraneoplastic diabetes. Nat Rev Gastroenterol Hepatol 2013;10:423–433.

18. Huxley R, Ansary-Moghaddam A, Berrington De-González A, et al. Type-II diabetes and pancreatic cancer: a meta-analysis of 36 studies. Br J Cancer 2005;92:2076–2083.

19. Chari ST, Leibson CL, Rabe KG, et al. Probability of pancreatic cancer following diabetes: a population-based study. Gastroenterology 2005;129:504–511.

20. National Cancer Institute. Could a diabetes diagnosis help detect pancreatic cancer early? 2021. http://www.cancer.gov/news-events/cancer-currents-blog/2021/pancreatic-cancer-diabetes-early-detection.

21. Singhi AD, Koay EJ, Chari ST, et al. Early detection of pancreatic cancer: opportunities and challenges. Gastroenterology 2019;156:2024–2040.

22. Lakhtakia S. Complications of diagnostic and therapeutic endoscopic ultrasound. Best Pract Res Clin Gastroenterol 2016;30:807–823.

23. McAllister F, Montiel MF, Uberoi GS, Uberoi AS, Maitra A, Bhutani MS. Current status and future directions for screening patients at high risk for pancreatic cancer. Gastroenterol Hepatol (N Y) 2017;13:268–275.

24. Canto MI, Almario JA, Schulick RD, et al. Risk of neoplastic progression in individuals at high risk for pancreatic cancer undergoing long-term surveillance. Gastroenterology 2018;155:740–751.e2.

25. National Comprehensive Cancer Network (NCCN). Clinical Practice Guidelines in Oncology. Pancreatic adenocarcinoma. V1.2021. https://www.nccn.org/guidelines/category_1

26. Marazzi I, Greenbaum BD, Low DHP, et al. Chromatin dependencies in cancer and inflammation. Nat Rev Mol Cell Biol 2018;19:245–261.

27. Feinberg AP. Epigenetic stochasticity, nuclear structure and cancer: the implications for medicine. J Intern Med 2014;276:5–11.

28. Beltrán-García J, Osca-Verdegal R, Mena-Mollá S, et al. Epigenetic IVD tests for personalized precision medicine in cancer. Front Genet 2019;10:621.

29. Tahiliani M, Koh KP, Shen Y, et al. Conversion of 5-methylcytosine to 5-hydroxymethylcytosine in mammalian DNA by MLL partner TET1. Science 2009;324:930–935.

30. Szulwach KE, Li X, Li Y, et al. Integrating 5-hydroxymethylcytosine into the epigenomic landscape of human embryonic stem cells. PLoS Genet 2011;7:e1002154.

31. Cui XL, Nie J, Ku J, et al. A human tissue map of 5-hydroxymethylcytosines exhibits tissue specificity through gene and enhancer modulation. Nat Commun 2020;11:6161.

32. Yang X, Han H, De Carvalho DD, et al. Gene body methylation can alter gene expression and is a therapeutic target in cancer. Cancer Cell 2014;26:577–590.

33. Nestor CE, Lentini A, Hägg Nilsson C, et al. 5-Hydroxymethylcytosine remodeling precedes lineage specification during differentiation of human CD4(+) T cells. Cell Rep 2016;16:559–570.

34. Song CX, Yin S, Ma L, et al. 5-Hydroxymethylcytosine signatures in cell-free DNA provide information about tumor types and stages. Cell Res 2017;27:1231–1242.

35. Li W, Zhang X, Lu X, et al. 5-Hydroxymethylcytosine signatures in circulating cell-free DNA as diagnostic biomarkers for human cancers. Cell Res 2017;27:1243–1257.

36. Guler GD, Ning Y, Ku CJ, et al. Detection of early stage pancreatic cancer using 5-hydroxymethylcytosine signatures in circulating cell free DNA. Nat Commun 2020;11:5270.

37. Robinson MD, McCarthy DJ, Smyth GK (2010). “edgeR: a Bioconductor package for differential expression analysis of digital gene expression data.” Bioinformatics, 26(1), 139–140.

38. Benjamini, Y. & Hochberg, Y. Controlling the false discovery rate: a practical and powerful approach to multiple testing. J. R. Stat. Soc. Series B (Methodological) 57, 289–300 (1995).

39. Li J, Cao G, Ma Q, et al. The bidirectional interaction between pancreatic cancer and diabetes. World J Surg Oncol 2012;10:171.

40. Serrano J, Andersen DK, Forsmark CE, et al. Consortium for the study of chronic pancreatitis, diabetes, and pancreatic cancer: from concept to reality. Pancreas 2018;47:1208–1212.

41. Centers for Disease Control and Prevention. National diabetes statistics report: estimates of diabetes and its burden in the United States, 2020. https://www.cdc.gov/diabetes/data/statistics-report/index.html.

42. Lin J, Thompson TJ, Cheng YJ, et al. Projection of the future diabetes burden in the United States through 2060. Popul Health Metr 2018;16:9.

43. Allison JE, Tekawa IS, Ransom LJ, et al. A comparison of fecal occult-blood tests for colorectal-cancer screening. N Engl J Med 1996;334:155–159.

44. National Lung Screening Trial Research Team. The National Lung Screening Trial: overview and study design. Radiology 2011;258:243–253.

45. Hayes DF. Defining clinical utility of tumor biomarker tests: a clinician’ s viewpoint. J Clin Oncol 2021;39:238–248.

46. O’ Brien DP, Sandanayake NS, Jenkinson C, et al. Serum CA19-9 is significantly upregulated up to 2 years before diagnosis with pancreatic cancer: implications for early disease detection. Clin Cancer Res 2015;21:622–631.

47. Campbell PJ, Yachida S, Mudie LJ, et al. The patterns and dynamics of genomic instability in metastatic pancreatic cancer. Nature 2010;467:1109–1113.

48. Maitra A, Sharma A, Brand RE, et al. A prospective study to establish a new-onset diabetes cohort: from the consortium for the study of chronic pancreatitis, diabetes, and pancreatic cancer. Pancreas 2018;47:1244–1248.

49. Zhang J, Han X, Gao C, et al. 5-Hydroxymethylome in circulating cell-free DNA as a potential biomarker for non-small-cell lung cancer. Genomics Proteomics Bioinformatics 2018;16:187–199.

50. Jones S, Zhang X, Parsons DW, et al. Core signaling pathways in human pancreatic cancers revealed by global genomic analyses. Science 2008;321:1801–1806.

51. Waddell N, Pajic M, Patch AM, et al. Whole genomes redefine the mutational landscape of pancreatic cancer. Nature 2015;518:495–501.

52. Baek B, Lee H. Prediction of survival and recurrence in patients with pancreatic cancer by integrating multi-omics data. Sci Rep 2020;10:18951.

53. Yachida S, White CM, Naito Y, et al. Clinical significance of the genetic landscape of pancreatic cancer and implications for identification of potential long-term survivors. Clin Cancer Res 2012;18:6339–6347.

54. Baumgart S, Chen NM, Siveke JT, et al. Inflammation-induced NFATc1-STAT3 transcription complex promotes pancreatic cancer initiation by Kras G12D. Cancer Discov 2014;4:688–701.

55. Roe JS, Hwang CI, Somerville TDD, et al. Enhancer reprogramming promotes pancreatic cancer metastasis. Cell 2017;170:875–888.

56. Davenport T, Kalakota R. The potential for artificial intelligence in healthcare. Future Healthc J 2019;6:94–98.

57. Kenner B, Chari ST, Kelsen D, et al. Artificial intelligence and early detection of pancreatic cancer: 2020 summative review. Pancreas 2021;50:251–279.

58. Kenner BJ, Chari ST, Cleeter DF, et al. Early detection of sporadic pancreatic cancer: strategic map for innovation—a white paper. Pancreas 2015; 44: 686–692.

59. Sanchez-Vega F, Mina M, Armenia J, et al. Oncogenic signaling pathways in the cancer genome atlas. Cell 2018;173:321–337.

60. Bijlsma MF, Sadanandam A, Tan P, et al. Molecular subtypes in cancers of the gastrointestinal tract. Nat Rev Gastroenterol Hepatol 2017;14:333–342.

61. Majumder S, Chari ST. Chronic pancreatitis. Lancet 2016;387:1957–1966.

62. Malka D, Hammel P, Maire F, et al. Risk of pancreatic adenocarcinoma in chronic pancreatitis. Gut 2002;51:849–852.

63. Surace AEA, Hedrich CM. The role of epigenetics in autoimmune/inflammatory disease. Front Immunol 2019;10:1525.

64. Kurkjian C, Kummar S, Murgo AJ. DNA methylation: its role in cancer development and therapy. Curr Probl Cancer 2008;32:187–235.

65. Liggett T, Melnikov A, Yi QL, et al. Differential methylation of cell-free circulating DNA among patients with pancreatic cancer versus chronic pancreatitis. Cancer 2010;116:1674–1680.

66. Grützmann R, Niedergethmann M, Pilarsky C, et al. Intraductal papillary mucinous tumors of the pancreas: biology, diagnosis, and treatment. Oncologist 2010;15:1294–1309.

67. Sato N, Fukushima N, Hruban RH, et al. CpG island methylation profile of pancreatic intraepithelial neoplasia. Mod Pathol 2008;21:238–244.

68. Tanaka M. Thirty years of experience with intraductal papillary mucinous neoplasm of the pancreas: from discovery to international consensus. Digestion 2014;90:265–272.

